# Reduced Likelihood of Hospitalization with the JN.1 or HV.1 SARS-CoV-2 Variants Compared to the EG.5 Variant

**DOI:** 10.1101/2024.05.08.24307003

**Authors:** Matthew E. Levy, Vanessa Chilunda, Richard E. Davis, Phillip R. Heaton, Pamala A. Pawloski, Jason D. Goldman, Cynthia A. Schandl, Lisa M. McEwen, Elizabeth T. Cirulli, Dana Wyman, Andrew Dei Rossi, Hang Dai, Magnus Isaksson, Nicole L. Washington, Tracy Basler, Kevin Tsan, Jason Nguyen, Jimmy Ramirez, Efren Sandoval, William Lee, James Lu, Shishi Luo

## Abstract

Within a multi-state viral genomic surveillance program, proportions of SARS-CoV-2 infections attributed to the JN.1 and HV.1 variants, compared to EG.5, were each lower among inpatients versus outpatients (aOR=0.33 [95% CI: 0.20-0.55] and aOR=0.62 [95% CI: 0.44-0.86], respectively). JN.1 and HV.1 variants may be associated with a lower risk of severe illness.

## INTRODUCTION

Throughout 2023, Omicron XBB-lineage variants were responsible for most SARS-CoV-2 infections in the U.S. [1]. Between January and May 2023, XBB.1.5 accounted for the majority of infections. This period was followed by multiple XBB variants and their sublineages cocirculating at lower levels. By August and October 2023, two sublineages of XBB.1.9.2 – EG.5 and its descendant HV.1 (see Supplementary Figure 1 for alternative pangolin designations) – each sequentially emerged as the most prevalent variants, respectively. By late December 2023, a descendant of BA.2.86, JN.1, became predominant. Compared to XBB variants, JN.1 possesses more than 30 mutations in the spike protein, including the notable L455S mutation, which may contribute to increased immune escape and infectivity [2,3].

The clinical severity of COVID-19 has differed across previous SARS-CoV-2 variants. On average, the Delta variant resulted in more severe illness than the Alpha variant [4,5], while the Omicron variant resulted in less severe illness than the Delta variant [6,7]. Among early Omicron sublineages, findings have been mixed, with some studies indicating more severe illness with BA.5 versus BA.1 or BA.2 variants and others reporting no difference [8,9]. Little is known regarding the relative severity of XBB lineages. In one study, XBB.1.16 was associated with a greater risk of severe outcomes compared to XBB.1.5 and XBB.1.9 [10]. Empirical data comparing severity between JN.1 and XBB-lineage variants is also scarce. One study suggests that severity is attenuated with BA.2.86 lineages (predominantly JN.1) [11].

Within a multi-state respiratory virus genomic surveillance program, we assessed whether the proportions of SARS-CoV-2 infections attributed to EG.5, HV.1, and JN.1 variants during periods of cocirculation differed among patients in inpatient care settings (considered more severe) versus outpatient care settings (considered more mild). This investigation offers valuable insights into whether the clinical severity of illness differs among the two most recently predominant XBB-lineage variants and the currently predominant JN.1 variant.

## METHODS

Within a pan-respiratory virus genomic surveillance program, residual clinical samples from patients who tested positive for a respiratory virus (molecular or antigen) were obtained from three health systems spanning five U.S. states (Supplementary Table 1). This analysis included samples collected starting on 14 May 2023 (i.e., the first 2-week period with EG.5 detected) through 20 January 2024. Only SARS-CoV-2-positive samples that were initially collected during outpatient or inpatient visits were included. Samples from emergency departments or other/unknown visit types were excluded. Patients’ demographic characteristics and COVID-19 vaccination history were extracted from electronic health records (EHRs) and state vaccine registries. Study protocols were approved by institutional review boards (central or local).

Viral sequencing was performed by Helix using a hybridization-capture based assay (Twist Biosciences) and short-read genome sequencing technology (Illumina) [12]. SARS-CoV-2 was identified in samples with reads that aligned to the reference genome, and lineages were assigned using pangolin version 4.3.1. Further details are provided in the Supplementary Methods.

The clinical visit type associated with sample collection served as a surrogate measure of the severity of illness at time of testing. Inpatient visits represented more severe illness compared to outpatient visits. The reason for the visit and patients’ specific symptoms were not available for analysis. COVID-19 vaccination status was assigned using the date of the most recent dose received prior to the specimen collection date (Supplementary Methods).

Proportions of outpatients and inpatients with EG.5, HV.1, and JN.1 variants were compared and plotted over 2-week intervals. EG.5 proportions were plotted up to the time of ≥5% HV.1 prevalence, and HV.1 proportions were plotted up to the time of ≥5% JN.1 prevalence (since the rise of emerging variants, which would be newly included in denominators, could influence observed proportions for previous variants). In multivariable logistic regression, the odds of infection with each variant were compared between outpatients and inpatients, adjusting for collection date (natural cubic spline), health system and state of residence, age group, sex, race/ethnicity, and COVID-19 vaccination status. Three pairwise comparisons were performed among EG.5, HV.1, and JN.1 during their full respective periods of cocirculation. We implemented a Bonferroni correction, whereby p<0.0167 was considered statistically significant. In exploratory analyses, these variants were also compared to other cocirculating XBB variants. Analyses were performed using R version 4.2.3.

## RESULTS

Between 14 May 2023 and 20 January 2024, 6,654 of 10,118 total samples (65.8%) that were collected during outpatient or inpatient visits were successfully assigned a SARS-CoV-2 lineage. Of those, 1,089 samples (16.4%) were collected during inpatient visits, compared to 5,565 samples (83.6%) collected during outpatient visits. XBB-lineage variants accounted for ≥90% of samples through the 2-week period ending 25 November 2023. The two most highly prevalent XBB-lineage variants, EG.5 and HV.1, reached peak prevalences of 29.3% and 26.7% during the 2-week periods ending 30 September and 25 November 2023, respectively. The prevalence of JN.1 increased from 20.3% during the 2-week period ending 9 December 2023 to 72.2% during the 2-week period ending 20 January 2024. Temporal patterns in variant prevalence for EG.5, HV.1, and JN.1 are shown in Supplementary Figure 2.

Patient characteristics overall and by visit type are described in Supplementary Table 2. Median age was higher among inpatients (74 years; IQR: 60-83) than among outpatients (50 years; IQR: 30-67). Female sex was less likely among inpatients compared to outpatients (52.2% vs 60.3%), and fewer inpatients than outpatients had ever received a COVID-19 vaccine (63.9% vs 68.4%).

Prior to HV.1 emergence, EG.5 accounted for a similar proportion of SARS-CoV-2 infections among outpatient and inpatient visits (Figure 1A). However, once HV.1 emerged, HV.1 consistently accounted for a greater proportion of infections among outpatient vs inpatient visits, prior to JN.1 emergence (Figure 1B). Once JN.1 emerged, JN.1 also accounted for more infections among outpatient vs inpatient visits (Figure 1C).

**Figure 1.**
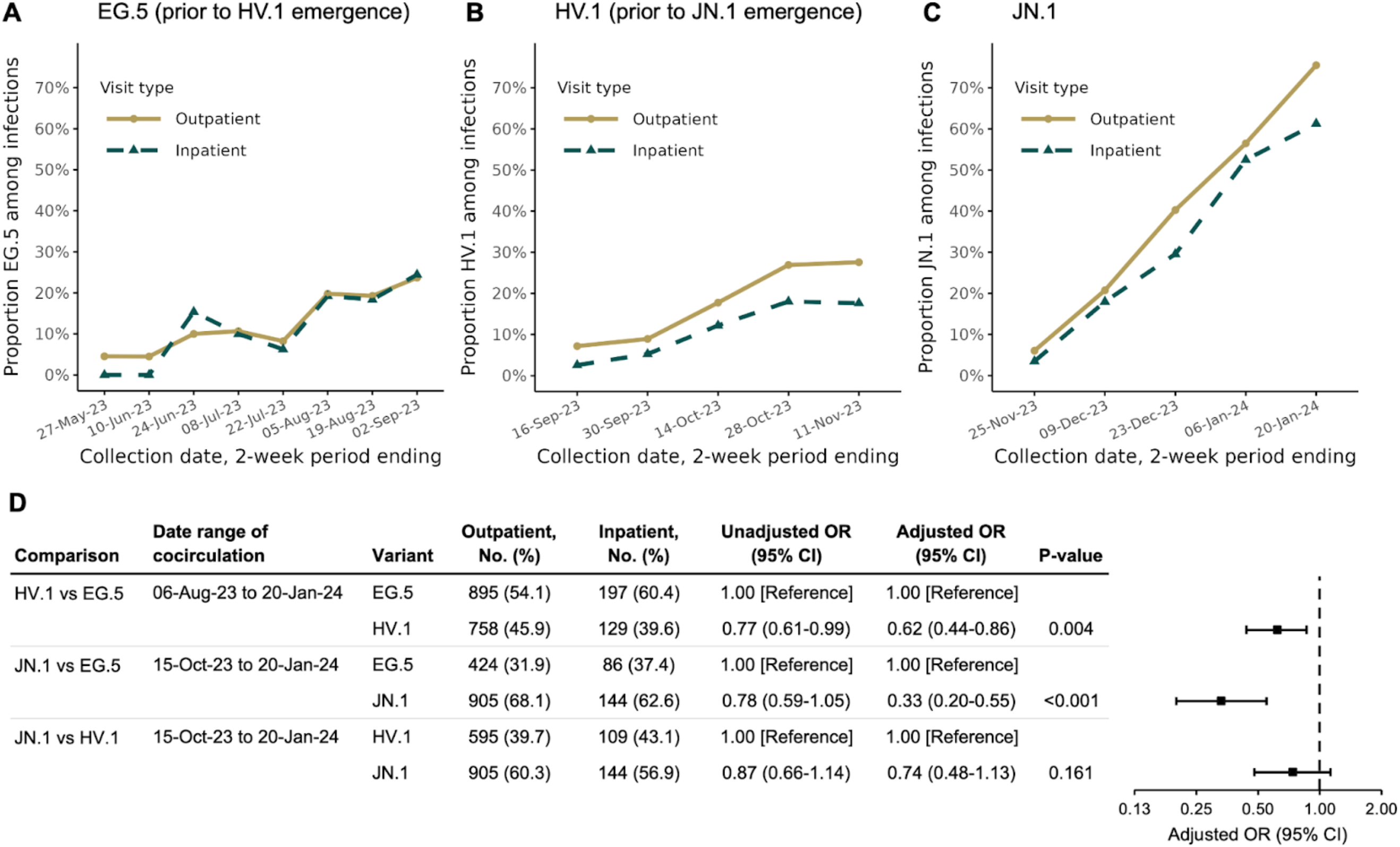
Biweekly prevalence of EG.5, HV.1, and JN.1 variants among samples collected during outpatient and inpatient visits and variant associations with inpatient visits. Proportions for EG.5 are shown during the period of EG.5 circulation up until the time of ≥5% HV.1 prevalence. Proportions for HV.1 are then shown up until the time of ≥5% JN.1 prevalence. Proportions for JN.1 are then shown through 20 January 2024. Pairwise variant associations were calculated among samples collected during all 2-week periods with variant cocirculation. Odds ratios (ORs) were adjusted for specimen collection date (as a natural cubic spline with degrees of freedom), health system and state of residence, age group (0-4 y, 5-11 y, 12-17 y, 18-49 y, 50-64 y, 65-74 y, 75-84 y, and ≥85 y), sex, race/ethnicity (Asian, non-Hispanic; Black, non-Hispanic; Hispanic; white, non-Hispanic; and other or unknown), and COVID-19 vaccination status (XBB.1.5-vaccinated, BA.4/BA.5-vaccinated, wild-type-vaccinated, and unvaccinated, based on the date of the most recent vaccine dose received prior to the sample collection date).

Both HV.1 and JN.1 were significantly less likely than EG.5 to account for infections among inpatients compared to outpatients, with adjusted odds ratios (aORs) of 0.62 (95% CI: 0.44-0.86, p=0.004) and 0.33 (95% CI: 0.20-0.55, p<0.001), respectively (Figure 1D). The distribution of JN.1 vs HV.1 did not show a significant difference between inpatient and outpatient visits (aOR=0.74; 95% CI: 0.48-1.13, p=0.16). Although precision was somewhat limited in additional exploratory comparisons, aOR point estimates for HV.1 and JN.1 were consistently <1 when compared to other cocirculating XBB-lineage variants, a trend not seen with EG.5 (Supplementary Figure 3).

## DISCUSSION

This study is among the first to provide empirical evidence regarding the relative severity of the currently predominant JN.1 variant and previously predominant XBB-lineage variants, which was enabled by the linkage of viral sequencing and patient-level EHR data. Among adults with medically attended SARS-CoV-2 infection, JN.1 and HV.1 infections were less prevalent among hospitalized patients and more prevalent among outpatients, compared to EG.5 infections during the same time period. This inverse association was nearly twice as strong for JN.1 than for HV.1, although the direct comparison of JN.1 versus HV.1 did not reach statistical significance. These variant differences were observed after adjusting for calendar time, demographics, and COVID-19 vaccination status. Our findings provide evidence that disease severity may have attenuated over the course of the 2023-2024 respiratory illness season, as the common circulating SARS-CoV-2 lineages have shifted from EG.5 to HV.1 to JN.1.

Our results are based on variant associations among infected individuals who accessed health system-administered SARS-CoV-2 testing. Healthcare-seeking and testing behaviors, which could vary by medical visit type and change over time, may have introduced differences between inpatients and outpatients beyond individuals’ disease severity and the patient characteristics measured. However, such behaviors are unlikely to correlate with one’s likelihood of infection with a specific variant over another at a given time point. Thus, primary findings are not likely to be explained by selection bias.

JN.1 has exhibited enhanced immune evasion and higher transmissibility compared to XBB variants, which might explain its rapid rise to predominance [2,3]. Although our findings suggest that JN.1 may be linked to less severe illness than EG.5, COVID-19 hospitalization rates could still be elevated during JN.1 predominance if the overall incidence of SARS-CoV-2 infection is higher. During the winter wave of 2023-2024, new COVID-19 hospital admissions reached their peak during the week ending 6 January 2024, just as JN.1 achieved predominance, which was then followed by a consistent decline [1]. It is uncertain whether this pattern can be partially attributed to JN.1 supplanting XBB lineages or if it was primarily driven by broader seasonal or temporal SARS-CoV-2 patterns.

Study limitations include the absence of data on symptoms, reasons for visits, and subsequent clinical outcomes. We also lacked information on social factors and underlying comorbid conditions. Visit type was determined at the time of sample collection for SARS-CoV-2 testing, and it is unknown whether outpatients were later hospitalized. Potential misclassification is likely to be non-differential with respect to variant. The precision of estimates was reduced when comparing JN.1, HV.1, and EG.5 to other cocirculating variants due to smaller sample sizes.

While an inverse association for JN.1 and HV.1 with hospitalization is reassuring, ongoing surveillance and further studies comparing patient outcomes across variants are essential. This study highlights the importance of timely linkage of viral genomic surveillance data with clinical records for monitoring outcomes associated with emerging SARS-CoV-2 variants.

## Data Availability

Data collected for this study are not available. Data sharing agreements between Helix and partner institutions prohibit Helix from making this dataset publicly available.

## NOTES

### Author contributions

All authors contributed substantively to this manuscript in the following ways: conceptualization (MEL, VC, SL), investigation (MEL, VC, RED, PRH, PAP, JDG, CAS, LMM, DW, ADR, HD, MI, NLW, TB, KT, JN, JR, ES, SL), data curation (MEL, VC, LMM, DW, ADR, HD, MI, NLW, SL), formal analysis (MEL), project administration (RED, PRH, PAP, JDG, CAS, ETC, NLW, WL, JL, SL), writing - original draft (MEL), and writing - review and editing (MEL, VC, RED, PRH, PAP, JDG, CAS, SL).

## Acknowledgements

We acknowledge the Helix Clinical Informatics and Bioinformatics teams for their contributions to electronic health record data and viral sequencing pipelines. We also acknowledge Catherine Clinton for her oversight and guidance in ensuring research compliance. We thank the investigators and staff at HealthPartners, Providence Health, and the Medical University of South Carolina who contributed to the ViEW Network™.

