## Supplementary Material for "Reduced Likelihood of Hospitalization with the JN.1 or HV.1 SARS-CoV-2 Variants Compared to the EG.5 Variant"

### Supplementary Methods

#### Viral sequencing

RNA was extracted from 400 µL of remnant patient anterior nares samples using the MagMAX Viral/Pathogen kit (ThermoScientific). RNA libraries (5µL RNA input volume) were generated using the Rapid RNA Library Kit protocol (Swift Biosciences/Integrated DNA Technologies). SARS-CoV-2 genome was enriched and captured using the Respiratory Virus Research Panel hybridization probe panel (Twist Biosciences). Samples were sequenced using the NovaSeq 6000 Sequencing system S1 flow cell, with S1 Reagent Kit v1.5 (300 cycles).

#### Bioinformatic analyses

The flow cell output was demultiplexed with bcl2fastq (Illumina) into per-sample FASTQ sequences. These sequences were then processed using the Helix fastagenerator pipeline to produce a consensus sequence FASTA file. First, reads were aligned to a reference set consisting of a representative genome of each respiratory virus that was targeted by the hybridization probes, and the human transcriptome (GENCODE v37) using BWA-MEM. The non-human reads were then re-aligned to an expanded set of respiratory viruses reference genomes including the SARS-CoV-2 genome (NCBI accession NC_045512.2) using a de Bruijn graph based algorithm (based on https://doi.org/10.1093/bioinformatics/btz814) to identify sequences that best match the reads. SARS-CoV-2 aligned reads were then marked for duplicates followed by variant calling using the Haplotyper algorithm (Sentieon, Inc). The consensus sequence for each sample was generated from the alignment (BAM) and variant call format (VCF) files according to the following criteria: coverage from at least 5 unique reads with at least 80% of the reads supporting the call at the base position. For the bases that did not meet this criteria, an N was reported. A sequence is considered to pass quality criteria if it contains at most 30% N bases.

### SARS-CoV-2 lineage designation

Viral sequences were assigned a PANGO lineage using pangolin (https://github.com/cov-lineages/pangolin). For this analysis, Pangolin data version v1.23.1 with Pangolin software v4.3.1 was used.

### COVID-19 vaccination status

COVID-19 vaccination status was assigned using the date of the most recent dose received prior to the specimen collection date, as documented in electronic health records and state public health vaccine registries (see Supplementary Table 1 for a description of each health system's vaccine data sources). Patients were classified as: 1) vaccinated with an XBB.1.5-adapted monovalent vaccine if their last dose occurred on/after 12 September 2023; 2) vaccinated with a BA.4/BA.5-adapted bivalent vaccine if their last dose occurred between 1 September 1 2022 and 11 September 2023; 3) vaccinated with an original wild-type monovalent vaccine if their last dose occurred before 1 September 2022, and 4) unvaccinated if they had received no prior COVID-19 vaccine dose.

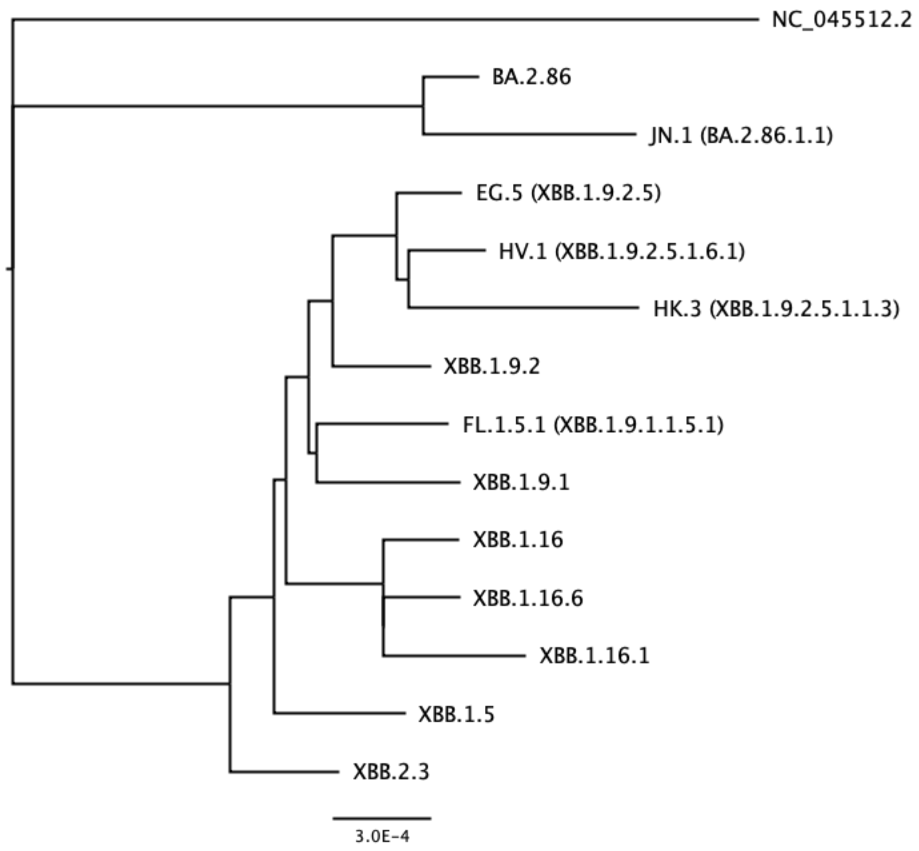


**Supplementary Figure 1. Phylogram of SARS-CoV-2 lineage categories.** The lengths of the branches are proportional to the genetic difference between variants. The distance bar shown represents 3 mutations per 10,000 bases. Except for FL.1.5.1, sublineages of XBB.1.9.1 are aggregated with XBB.1.9.1. Except for HV.1 and HK.3, sublineages of EG.5 are aggregated with EG.5. Except for EG.5, HV.1, and HK.3, sublineages of XBB.1.9.2 are aggregated with XBB.1.9.2. Except for XBB.1.16.1 and XBB.1.16.6, sublineages of XBB.1.16 are aggregated with XBB.1.16. Except for JN.1, sublineages of BA.2.86 are aggregated with BA.2.86. Sublineages of each other named lineage are aggregated with the respective lineage. NC_045512.2 is the accession number for the originally published sequence for SARS-CoV-2, representing PANGO lineage “B”.

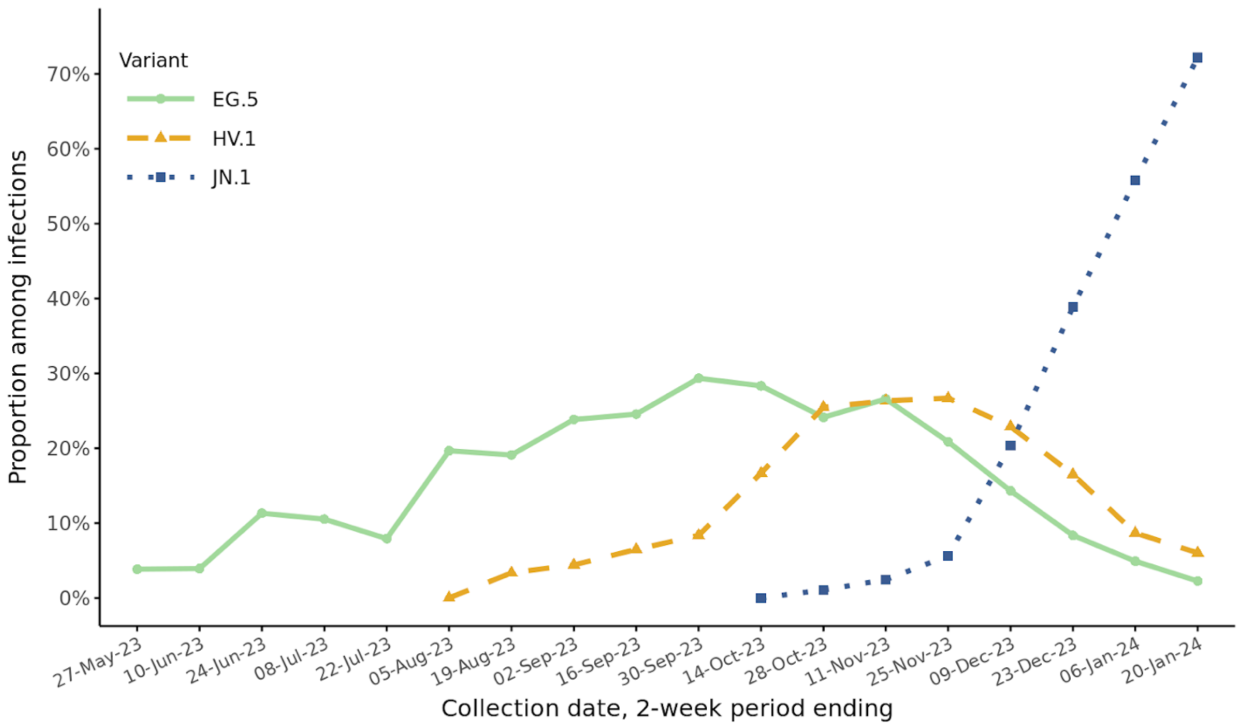


**Supplementary Figure 2. Biweekly prevalence of EG.5, HV.1, and JN.1 variants between 14 May 2023 and 20 January 2024 in the ViEW Network multi-state viral genomic surveillance program.**

| Comparison | Date range of cocirculation | Variant | Outpatient, No. (%) | Inpatient, No. (%) | Unadjusted OR (95% CI) | Adjusted OR (95% CI) |
|---|---|---|---|---|---|---|
| EG.5 vs XBB.1.16.1 | 14-May-23 to 20-Jan-24 | XBB.1.16.1 | 536 (35.9) | 107 (33.5) | 1.00 [Reference] | 1.00 [Reference] |
| | | EG.5 | 958 (64.1) | 212 (66.5) | 1.11 (0.86-1.43) | 1.10 (0.80-1.50) |
| EG.5 vs XBB.1.5 | 14-May-23 to 20-Jan-24 | XBB.1.5 | 529 (35.6) | 105 (33.1) | 1.00 [Reference] | 1.00 [Reference] |
| | | EG.5 | 958 (64.4) | 212 (66.9) | 1.11 (0.86-1.44) | 1.38 (0.97-1.98) |
| EG.5 vs XBB.2.3 | 14-May-23 to 20-Jan-24 | XBB.2.3 | 400 (29.5) | 79 (27.1) | 1.00 [Reference] | 1.00 [Reference] |
| | | EG.5 | 958 (70.5) | 212 (72.9) | 1.12 (0.84-1.49) | 1.05 (0.73-1.52) |
| EG.5 vs XBB.1.16.6 | 14-May-23 to 20-Jan-24 | XBB.1.16.6 | 296 (23.6) | 66 (23.7) | 1.00 [Reference] | 1.00 [Reference] |
| | | EG.5 | 958 (76.4) | 212 (76.3) | 0.99 (0.73-1.35) | 1.48 (1.00-2.18) |
| EG.5 vs FL.1.5.1 | 09-Jul-23 to 20-Jan-24 | FL.1.5.1 | 220 (19.0) | 44 (17.5) | 1.00 [Reference] | 1.00 [Reference] |
| | | EG.5 | 938 (81.0) | 208 (82.5) | 1.11 (0.78-1.58) | 1.27 (0.81-1.99) |
| HV.1 vs XBB.1.16.1 | 06-Aug-23 to 20-Jan-24 | XBB.1.16.1 | 510 (40.2) | 101 (43.9) | 1.00 [Reference] | 1.00 [Reference] |
| | | HV.1 | 758 (59.8) | 129 (56.1) | 0.86 (0.65-1.14) | 0.66 (0.45-0.96) |
| HV.1 vs XBB.1.5 | 06-Aug-23 to 20-Jan-24 | XBB.1.5 | 369 (32.7) | 68 (34.5) | 1.00 [Reference] | 1.00 [Reference] |
| | | HV.1 | 758 (67.3) | 129 (65.5) | 0.92 (0.67-1.27) | 0.72 (0.45-1.16) |
| HV.1 vs XBB.2.3 | 06-Aug-23 to 20-Jan-24 | XBB.2.3 | 329 (30.3) | 63 (32.8) | 1.00 [Reference] | 1.00 [Reference] |
| | | HV.1 | 758 (69.7) | 129 (67.2) | 0.89 (0.64-1.23) | 0.61 (0.39-0.98) |
| HV.1 vs XBB.1.16.6 | 06-Aug-23 to 20-Jan-24 | XBB.1.16.6 | 276 (26.7) | 61 (32.1) | 1.00 [Reference] | 1.00 [Reference] |
| | | HV.1 | 758 (73.3) | 129 (67.9) | 0.77 (0.55-1.08) | 0.90 (0.56-1.46) |
| HV.1 vs FL.1.5.1 | 06-Aug-23 to 20-Jan-24 | FL.1.5.1 | 215 (22.1) | 42 (24.6) | 1.00 [Reference] | 1.00 [Reference] |
| | | HV.1 | 758 (77.9) | 129 (75.4) | 0.87 (0.60-1.27) | 0.79 (0.46-1.35) |
| JN.1 vs XBB.1.16.1 | 15-Oct-23 to 20-Jan-24 | XBB.1.16.1 | 218 (19.4) | 43 (23.0) | 1.00 [Reference] | 1.00 [Reference] |
| | | JN.1 | 905 (80.6) | 144 (77.0) | 0.81 (0.56-1.17) | 0.89 (0.51-1.53) |
| JN.1 vs XBB.1.5 | 15-Oct-23 to 20-Jan-24 | XBB.1.5 | 163 (15.3) | 36 (20.0) | 1.00 [Reference] | 1.00 [Reference] |
| | | JN.1 | 905 (84.7) | 144 (80.0) | 0.72 (0.48-1.08) | 0.65 (0.34-1.24) |
| JN.1 vs XBB.2.3 | 15-Oct-23 to 20-Jan-24 | XBB.2.3 | 187 (17.1) | 21 (12.7) | 1.00 [Reference] | 1.00 [Reference] |
| | | JN.1 | 905 (82.9) | 144 (87.3) | 1.42 (0.87-2.30) | 0.66 (0.35-1.24) |

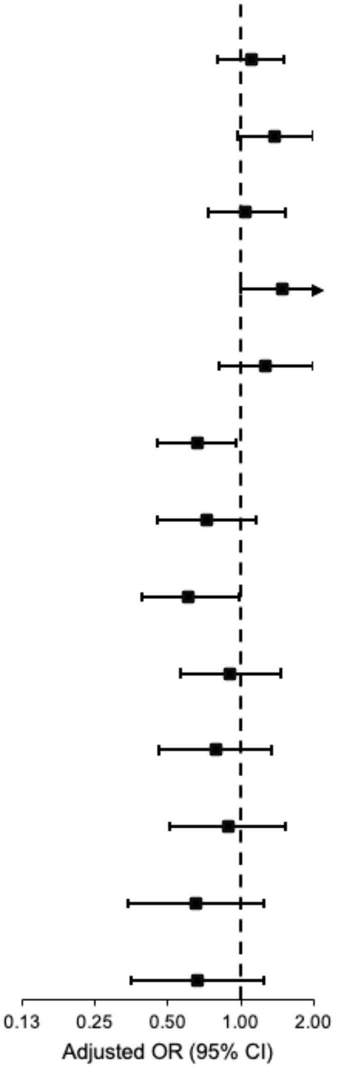


**Supplementary Figure 3. Variant associations with inpatient visits for EG.5, HV.1, and JN.1 variants compared to other circulating variants.** Pairwise variant associations were calculated among samples collected during all 2-week periods with variant cocirculation. Odds ratios (ORs) were adjusted for specimen collection date (as a natural cubic spline with 4 degrees of freedom), health system and state of residence, age group (0-4 y, 5-11 y, 12-17 y, 18-49 y, 50-64 y, 65-74 y, 75-84 y, and ≥85 y), sex, race/ethnicity (Asian, non-Hispanic; Black, non-Hispanic; Hispanic; white, non-Hispanic; and other or unknown), and COVID-19 vaccination status (XBB.1.5-vaccinated, BA.4/BA.5-vaccinated, wild-type-vaccinated, and unvaccinated, based on the date of the most recent vaccine dose received prior to the sample collection date).

**Supplementary Table 1. Health Systems Participating in the ViEW Network multi-state viral genomic surveillance program.**

| Health system | HealthPartners | Providence Health | Medical University of South Carolina |
|---|---|---|---|
| **Protocol number** | 0006-001 | 0006-001 | 0006-001 |
| **Institutional Review Board (IRB)** | WCG IRB | WCG IRB | MUSC IRB |
| **IRB approval number** | 20224919 | 20224919 | Pro00129083 |
| **Geographic region** | Minnesota and western Wisconsin | Eastern Washington and southern California | South Carolina |
| **Date range of patients included** | May 14, 2023–January 20, 2024 | May 25, 2023–January 20, 2024 | August 31, 2023–January 13, 2024 |
| **Sample size** | 6,139 | 379 | 136 |
| **Vaccine record data sources** | Epic electronic health record, data linkage with the Minnesota Immunization Information Connection (MIIC), and medical and pharmacy claims data | Epic electronic health record and data linkages with the California Immunization Registry (CAIR) and Washington State Immunization Information System (WAIIS) | Epic electronic health record; immunization records in the South Carolina Statewide Immunization Online Network (SIMON) are reconciled in Epic after a patient's appointment |
| **Informed consent** | Waiver of consent obtained | Waiver of consent obtained | Waiver of consent obtained |
| **Data privacy** | Limited dataset stripped of direct identifiers | Limited dataset stripped of direct identifiers | Limited dataset stripped of direct identifiers |

**Supplementary Table 2. Patient Characteristics Overall and Stratified by Visit Type.**

| Characteristic | Overall (n=6,654), No. (%) | Visit type: Outpatient (n=5,565), No. (%) | Visit type: Inpatient (n=1,089), No. (%) |
|---|---|---|---|
| Health system | | | |
| HealthPartners | 6,139 (92.3) | 5,425 (97.5) | 714 (65.6) |
| Medical University of South Carolina | 136 (2.0) | 40 (0.7) | 96 (8.8) |
| Providence Health and Services | 379 (5.7) | 100 (1.8) | 279 (25.6) |
| Age group | | | |
| 0 to 4 y | 376 (5.7) | 351 (6.3) | 25 (2.3) |
| 5 to 11 y | 141 (2.1) | 139 (2.5) | 2 (0.2) |
| 12 to 17 y | 192 (2.9) | 184 (3.3) | 8 (0.7) |
| 18 to 49 y | 2,261 (34.0) | 2,108 (37.9) | 153 (14.0) |
| 50 to 64 y | 1,321 (19.9) | 1,177 (21.2) | 144 (13.2) |
| 65 to 74 y | 1,082 (16.3) | 859 (15.4) | 223 (20.5) |
| 75 to 84 y | 881 (13.2) | 572 (10.3) | 309 (28.4) |
| ≥85 y | 400 (6.0) | 175 (3.1) | 225 (20.7) |
| Female | 3,927 (59.0) | 3,358 (60.3) | 569 (52.2) |
| Race and ethnicity | | | |
| Asian, non-Hispanic | 412 (6.2) | 374 (6.7) | 38 (3.5) |
| Black, non-Hispanic | 809 (12.2) | 699 (12.6) | 110 (10.1) |
| Hispanic | 312 (4.7) | 284 (5.1) | 28 (2.6) |
| White, non-Hispanic | 4,147 (62.3) | 3,530 (63.4) | 617 (56.7) |
| Other or unknown | 974 (14.6) | 678 (12.2) | 296 (27.2) |
| COVID-19 vaccination status[a] | | | |
| Unvaccinated | 2,150 (32.3) | 1,757 (31.6) | 393 (36.1) |
| Vaccinated with a wild-type vaccine | 2,861 (43.0) | 2,438 (43.8) | 423 (38.8) |
| Vaccinated with a BA.4/BA.5 vaccine | 1,139 (17.1) | 929 (16.7) | 210 (19.3) |
| Vaccinated with an XBB.1.5 vaccine | 504 (7.6) | 441 (7.9) | 63 (5.8) |
| SARS-CoV-2 variant[b] | | | |
| EG.5 | 1,170 (17.6) | 958 (17.2) | 212 (19.5) |
| JN.1 | 1,049 (15.8) | 905 (16.3) | 144 (13.2) |
| HV.1 | 887 (13.3) | 758 (13.6) | 129 (11.8) |

| | | | |
|---|---|---|---|
| XBB.1.16.1 | 643 (9.7) | 536 (9.6) | 107 (9.8) |
| XBB.1.5 | 634 (9.5) | 529 (9.5) | 105 (9.6) |
| XBB.2.3 | 479 (7.2) | 400 (7.2) | 79 (7.3) |
| XBB.1.16.6 | 362 (5.4) | 296 (5.3) | 66 (6.1) |
| FL.1.5.1 | 264 (4.0) | 220 (4.0) | 44 (4.0) |
| HK.3 | 231 (3.5) | 186 (3.3) | 45 (4.1) |
| XBB.1.16 | 236 (3.5) | 185 (3.3) | 51 (4.7) |
| BA.2.86 | 148 (2.2) | 126 (2.3) | 22 (2.0) |
| XBB.1.9.1 | 139 (2.1) | 122 (2.2) | 17 (1.6) |
| XBB.1.9.2 | 86 (1.3) | 67 (1.2) | 19 (1.7) |
| Other XBB | 230 (3.5) | 192 (3.5) | 38 (3.5) |
| Other non-XBB | 96 (1.4) | 85 (1.5) | 11 (1.0) |

[a] Defined by whether there was ≥1 COVID-19 vaccine record prior to the specimen collection date and by the date of the most recent dose received.

[b] SARS-CoV-2 variant lineages were assigned using pangolin version 4.3.1. Except for HV.1 and HK.3, sublineages of EG.5 are aggregated with EG.5. Except for XBB.1.16.1 and XBB.1.16.6, sublineages of XBB.1.16 are aggregated with XBB.1.16. Except for JN.1, sublineages of BA.2.86 are aggregated with BA.2.86. Except for FL.1.5.1, sublineages of XBB.1.9.1 are aggregated with XBB.1.9.1. Except for EG.5, HV.1, and HK.3, sublineages of XBB.1.9.2 are aggregated with XBB.1.9.2. Sublineages of each other named lineage are aggregated with the respective lineage. JN.1 is also known as BA.2.86.1.1; HV.1 as XBB.1.9.2.5.1.6.1; EG.5 as XBB.1.9.2.5; HK.3 as XBB.1.9.2.5.1.1.3; and FL.1.5.1 as XBB.1.9.1.1.5.1.
